# Altered sleep architecture in diabetes and prediabetes: findings from the Baependi Heart Study

**DOI:** 10.1101/2023.03.23.23287631

**Authors:** Daniel M. Chen, Tâmara P. Taporoski, Shaina J. Alexandria, David A. Aaby, Felipe Beijamini, Jose E. Krieger, Malcolm von Schantz, Alexandre Pereira, Kristen L. Knutson

## Abstract

**Objective:** People with diabetes are more likely to have obstructive sleep apnea, but there are few studies examining sleep architecture in people with diabetes, especially in the absence of moderate-severe sleep apnea. Therefore, we compared sleep architecture among people with diabetes, prediabetes or neither condition, whilst excluding people with moderate-severe sleep apnea.

**Research design and methods:** This sample is from the Baependi Heart Study, a prospective, family-based cohort of adults in Brazil. 1,074 participants underwent at-home polysomnography (PSG). Diabetes was defined as 1) FBG>125 OR 2) HbA1c>6.4 OR 3) taking diabetic medication, and prediabetes was defined as 1) [(5.7≤HbA1c≤6.4) OR (100≤FBG≤125)] AND 2) not taking diabetic medication. We excluded participants that had an apnea-hypopnea index (AHI)>30 from these analyses to reduce confounding due to severe sleep apnea. We compared sleep stages among the 3 groups.

**Results:** Compared to those without diabetes, we found shorter REM duration for participants with diabetes (−6.7min, 95%CI -13.2, -0.1) or prediabetes (−5.9min, 95%CI -10.5, -1.3), even after adjusting for age, gender, BMI, and AHI. Diabetes was also associated with lower total sleep time (−13.7min, 95%CI -26.8, -0.6), longer slow-wave sleep (N3) duration (+7.6min, 95%CI 0.6, 14.6) and higher N3 percentage (+2.4%, 95%CI 0.6, 4.2), compared to those without diabetes.

**Conclusions:** People with diabetes and prediabetes had less REM sleep after taking into account potential confounders, including AHI. People with diabetes also had more N3 sleep. These results suggest that diabetes is associated with different sleep architecture, even in the absence of moderate-severe sleep apnea.

---

Type 2 diabetes mellitus is one of the most common chronic medical conditions in the world, affecting approximately 537 million people worldwide in 2021 (1). It can lead to medical complications and early mortality, and reduced quality of life. Individuals with diabetes often report insufficient sleep duration or decreased sleep quality (2; 3). Type 2 diabetes is also often associated with sleep disordered breathing, particularly obstructive sleep apnea (OSA) (4; 5). Because of its association with glucose homeostasis, healthy sleep may be especially important in type 2 diabetes. For example, studies have observed associations between poor subjective sleep and higher HbA1c (6-9). Fewer studies have examined objectively assessed sleep architecture via polysomnography in people with diabetes and prediabetes.

Polysomnography (PSG) provides detailed information on the distribution of time spent in each sleep stage across the night, known as sleep architecture. The two primary stages of sleep are rapid-eye movement (REM) and non-REM (NREM) sleep; N3 is the deepest stage of NREM. Alterations in sleep architecture have also been shown to cause impaired metabolic function. For example, two experimental studies that suppressed N3 in young healthy adults observed impairments in glucose metabolism and insulin sensitivity (10; 11). In addition, experimental sleep restriction which resulted in less REM sleep was associated with positive energy balance (12), which could increase risk of weight gain. Therefore, sleep architecture may be particularly important for people with diabetes since they rely so heavily on tight glycemic and weight control to avoid complications. A few studies have investigated whether sleep architecture is disturbed in type 2 diabetes, but results are conflicting (13-16). These studies varied in sample sizes, most had fewer than 200 participants, and few accounted for OSA. OSA is commonly associated with disturbed sleep architecture, including decreased N3 and REM duration (17). Therefore, it is unclear whether type 2 diabetes is associated with poorer sleep architecture, such as less N3 or REM sleep, particularly in the absence of moderate-severe sleep apnea. The goal of this study was to compare sleep architecture, accounting for OSA, among those with diabetes, prediabetes and neither condition in a large, observational study from Brazil. We hypothesized that people with prediabetes and diabetes will have less N3 and REM sleep than those with neither condition.

## Research Design and Methods

### Sample

This sample is from an ancillary study to the Baependi Heart Study (BHS), a family-based cohort of adults in a rural town in the state of Minas Gerais in southeastern Brazil. The recruitment methodology for the BHS has been described previously (18). Briefly, adult probands were selected from the community at random across 11 out of the 12 census districts in Baependi. Once probands were enrolled, all of their first, second and third degree relatives, who were at least 18 years old, were invited to participate. Since 2019, BHS participants became eligible to participate in an ancillary study to collect full ambulatory polysomnography (PSG) in the cohort. Collection and processing of PSG data from the BHS is ongoing. The BHS subsample used herein includes participants whose PSG data was collected and processed as of November 10, 2022.

### Measures

Participants underwent one night of full ambulatory polysomnography (PSG) and provided blood samples before and after sleep. The PSG recording included the following channels: EEG, EOG, EMG, nasal airflow, respiratory effort from two RIP belts, and pulse oximetry. The PSG recordings were staged and scored by qualified polysomnographic technologists using the American Academy of Sleep Medicine (AASM) scoring criteria (19). The scored PSG data was used to calculate the following characteristics of sleep: total sleep time (TST; hours), total time in NREM stage 2 (N2; minutes), total time in NREM stage 3 (N3; minutes), total time in REM sleep (minutes), and total wake after sleep onset (WASO; minutes). We also calculated the percentages of total sleep time spent in N3 and REM. Finally, we calculated the apnea-hypopnea index (AHI) as an indicator of sleep disordered breathing. The AHI was calculated as the average number of apneas and hypopneas that occur per hour of sleep.

We used the fasting blood sample collected in the morning to assay both fasting blood glucose (FBG) and HbA1c levels. We also asked participants to report their medications. Using this information, we classified participants into three diabetes groups: diabetes, prediabetes, or neither condition. Diabetes was defined as meeting at least one of the following criteria: 1) an FBG of >125 mg/dL, 2) an HbA1c > 6.4, or 3) taking diabetic medication. We defined prediabetes as: 1) not in the diabetes category, 2) HbA1c ≥5.7 and ≤ 6.4 or fasting glucose ≥100 and ≤125 mg/dl, and 3) not taking diabetic medication. All other individuals were classified as having neither condition.

### Statistical Analysis

All statistical analyses were conducted using R Statistical Software version 4.1.2 (20). The BHS sleep ancillary sample as of November 11, 2022 included 1608 participants. Participants were excluded if the PSG recording was invalid due to poor quality (n=280), if the PSG recorded less than 4 hours of total sleep time (n=112), if systolic blood pressure < 50 mmHg or diastolic blood pressure < 30 mmHg (n=4), and if AHI ≥ 30 events/hour or AHI was missing (n=138). Our final analytic sample included 1074 participants.

Sample characteristics were summarized as either mean (standard deviation) or n (%), as appropriate. We used linear regression to model the association between each PSG sleep characteristic and diabetes groups (diabetes, prediabetes, neither condition). For each sleep outcome, we fit unadjusted models (Model 1) and fully adjusted models (Model 2) that adjusted for age, gender, body mass index (BMI), and AHI. We conducted sensitivity analyses using the same modeling procedure described above to assess whether results differed in a sample restricted to participants with moderate-severe sleep apnea (AHI < 15; N=890). Finally, we examined potential effect modification by age since those with prediabetes and diabetes tended to be older. To do this, we included age*diabetes interaction terms in the fully adjusted models. All significance tests were two sided, and statistical estimates are reported with accompanying 95% confidence intervals and p-values.

### Data Availability Statement

The datasets generated during and/or analyzed in the current study are available from the corresponding author upon reasonable request and establishment of a data use agreement.

### Ethical Approval

The Baependi Heart Study protocol conformed to the tenets of the Declaration of Helsinki and was approved by the Ethics Committee of the Hospital das Clínicas, University of São Paulo, Brazil (approval number 0494/10).

## Results

The characteristics of the 1,074 participants are summarized in **Table 1**. The majority of the participants were female (64.2%), and the percentage of females was similar across all three diabetes groups (prediabetes, diabetes, neither condition). Figure 1 presents boxplots of four PSG measures by diabetes group: TST, WASO, N3 and REM. In unadjusted models (Table 2), both prediabetes and diabetes were associated with shorter TST, more WASO, less REM and lower REM percentage. Prediabetes was also associated with less N3 and low N3 percentage in unadjusted models.

**Table 1:** Descriptive characteristics of the sample with AHI<30 events/hour.

| **Characteristic** | **Overall**<br>**(N = 1,074)** | **Neither condition**<br>**(N = 700)** | **Prediabetes**<br>**(N = 261)** | **Diabetes**<br>**(N = 113)** |
| --- | --- | --- | --- | --- |
| Age (y) | 48.8 ± 14.0 | 44.6 ± 12.8 | 55.7 ± 12.6 | 59.1 ± 12.1 |
| Gender (% male) | 35.8 | 37.1 | 34.5 | 31.0 |
| FBG (mg/dL) | 86 ± 22 | 80 ± 9 | 87 ± 12 | 121 ± 47 |
| HbA1c (%) | 5.60 ± 0.88 | 5.24 ± 0.30 | 5.86 ± 0.22 | 7.31 ± 1.82 |
| BMI | 26.7 ± 5.1 | 25.8 ± 4.6 | 28.0 ± 5.2 | 29.7 ± 6.1 |
| AHI | 6.9 ± 7.6 | 5.7 ± 7.1 | 8.7 ± 7.7 | 10.4 ± 8.8 |
| TST (h) | 6.6 ± 1.0 | 6.7 ± 1.0 | 6.5 ± 1.1 | 6.2 ± 1.0 |
| WASO (min) | 59 ± 41 | 54 ± 38 | 65 ± 43 | 73 ± 45 |
| N2 (min) | 225 ± 52 | 225 ± 50 | 226 ± 57 | 217 ± 57 |
| N3 (min) | 46.1 ± 34.8 | 48.5 ± 33.6 | 41.4 ± 35.8 | 42.5 ± 38.3 |
| N3 % | 11.7 ± 8.9 | 12.1 ± 8.4 | 10.8 ± 9.3 | 11.5 ± 10.4 |
| REM (min) | 79.8 ± 31.1 | 84.5 ± 30.2 | 71.9 ± 32.3 | 68.9 ± 29.0 |
| REM % | 20.0 ± 6.8 | 20.9 ± 6.3 | 18.3 ± 7.4 | 18.4 ± 7.3 |

All measurements reported as mean ± SD or percentage (%). Abbreviations are defined as
follows: FBG = fasting blood glucose, HbA1c = hemoglobin A1c, BMI = body mass index, AHI
= apnea-hypopnea index, TST = total sleep time, WASO = wake after sleep onset.

**Table 2.**
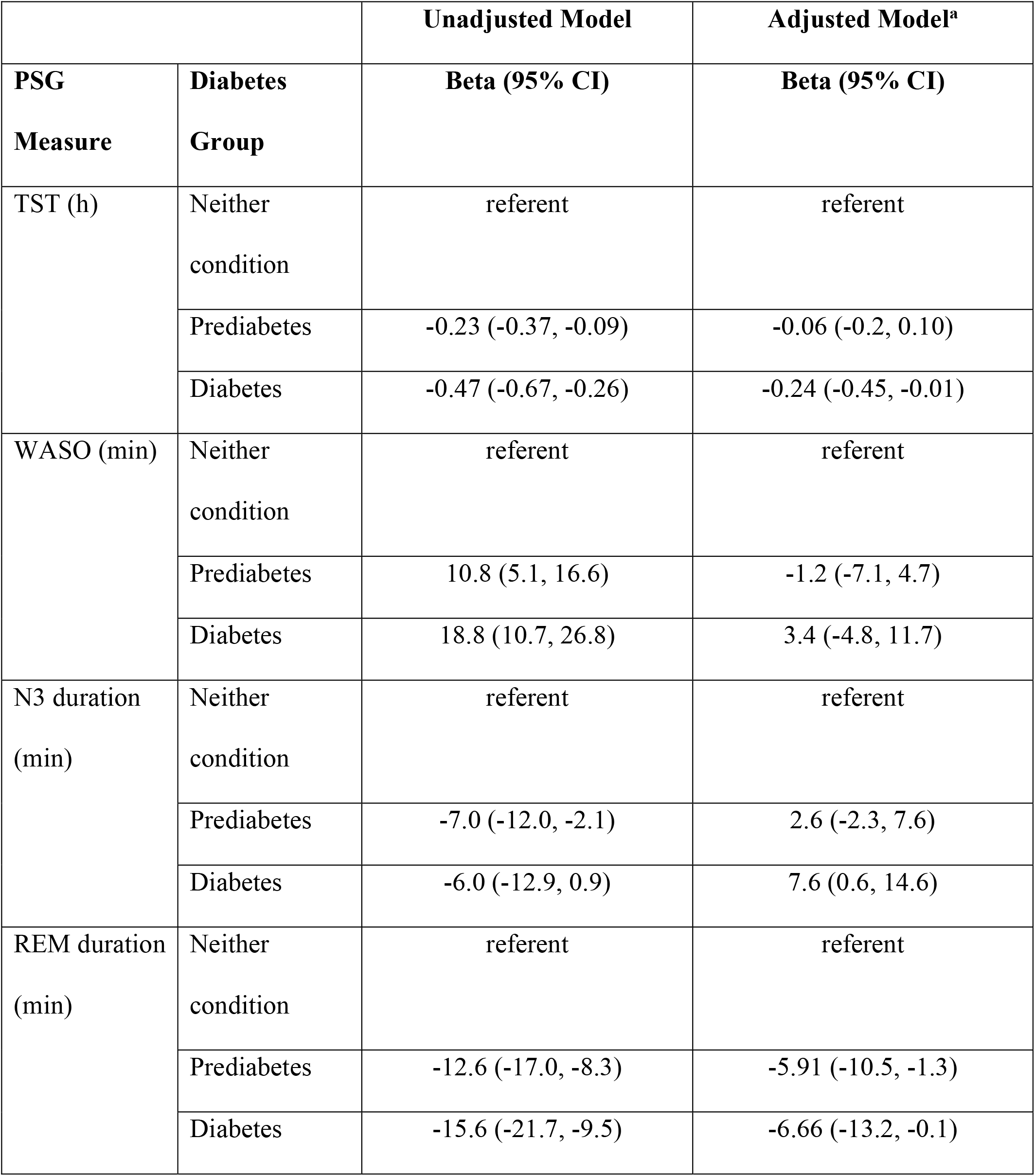
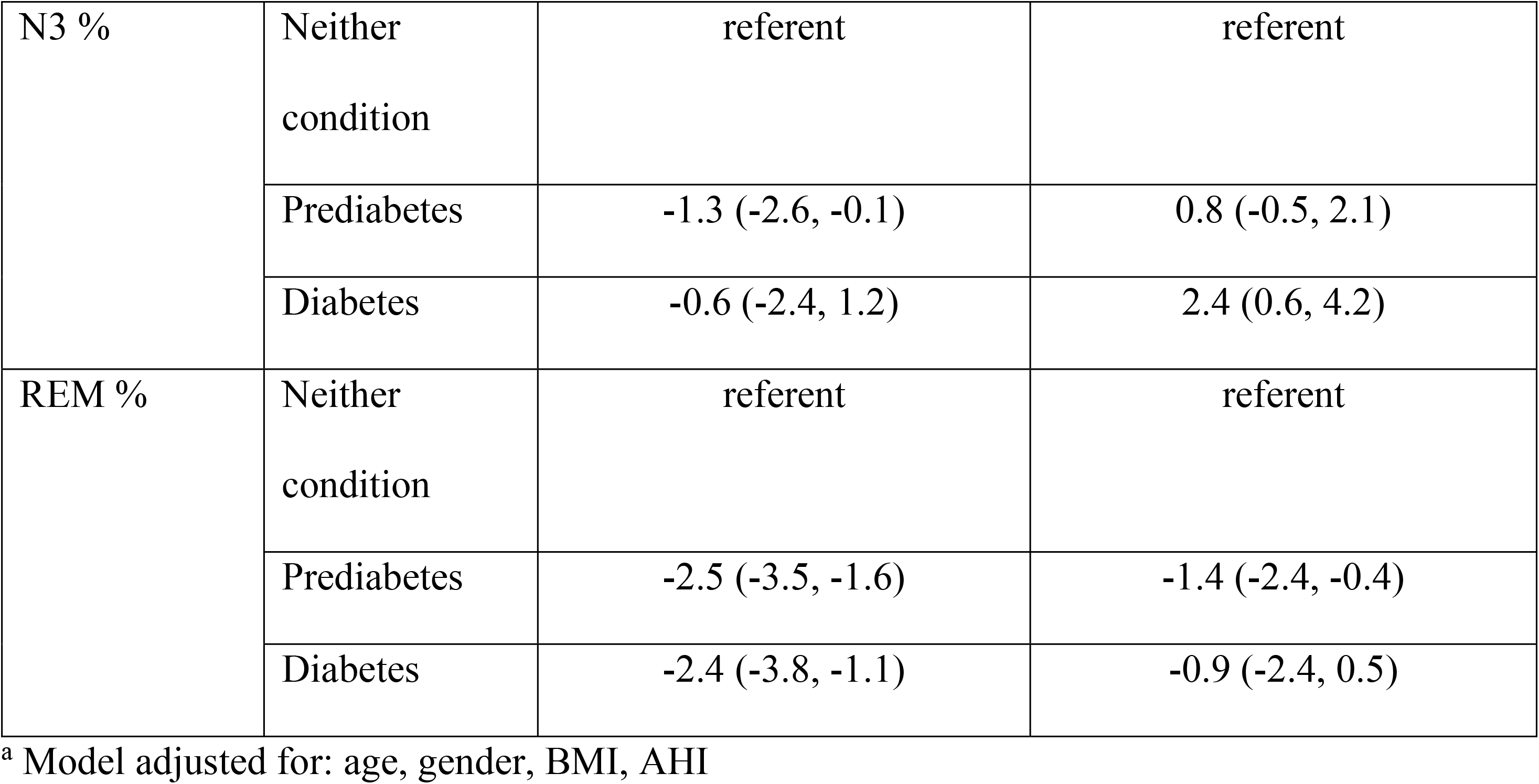
Regression models comparing PSG measures among individuals with neither condition, prediabetes and diabetes in sample with AHI<30 events/hour.

|  |  | Unadjusted Model | Adjusted Model<sup>a</sup> |
| --- | --- | --- | --- |
| PSG Measure | Diabetes Group | Beta (95% CI) | Beta (95% CI) |
| TST (h) | Neither condition | referent | referent |
|  | Prediabetes | -0.23 (-0.37, -0.09) | -0.06 (-0.2, 0.10) |
|  | Diabetes | -0.47 (-0.67, -0.26) | -0.24 (-0.45, -0.01) |
| WASO (min) | Neither condition | referent | referent |
|  | Prediabetes | 10.8 (5.1, 16.6) | -1.2 (-7.1, 4.7) |
|  | Diabetes | 18.8 (10.7, 26.8) | 3.4 (-4.8, 11.7) |
| N3 duration (min) | Neither condition | referent | referent |
|  | Prediabetes | -7.0 (-12.0, -2.1) | 2.6 (-2.3, 7.6) |
|  | Diabetes | -6.0 (-12.9, 0.9) | 7.6 (0.6, 14.6) |
| REM duration (min) | Neither condition | referent | referent |
|  | Prediabetes | -12.6 (-17.0, -8.3) | -5.91 (-10.5, -1.3) |
|  | Diabetes | -15.6 (-21.7, -9.5) | -6.66 (-13.2, -0.1) |
| N3 % | Neither<br>condition | referent | referent |
|  | Prediabetes | -1.3 (-2.6, -0.1) | 0.8 (-0.5, 2.1) |
|  | Diabetes | -0.6 (-2.4, 1.2) | 2.4 (0.6, 4.2) |
| REM % | Neither<br>condition | referent | referent |
|  | Prediabetes | -2.5 (-3.5, -1.6) | -1.4 (-2.4, -0.4) |
|  | Diabetes | -2.4 (-3.8, -1.1) | -0.9 (-2.4, 0.5) |

<sup>a</sup> Model adjusted for: age, gender, BMI, AHI

**Figure 1:**
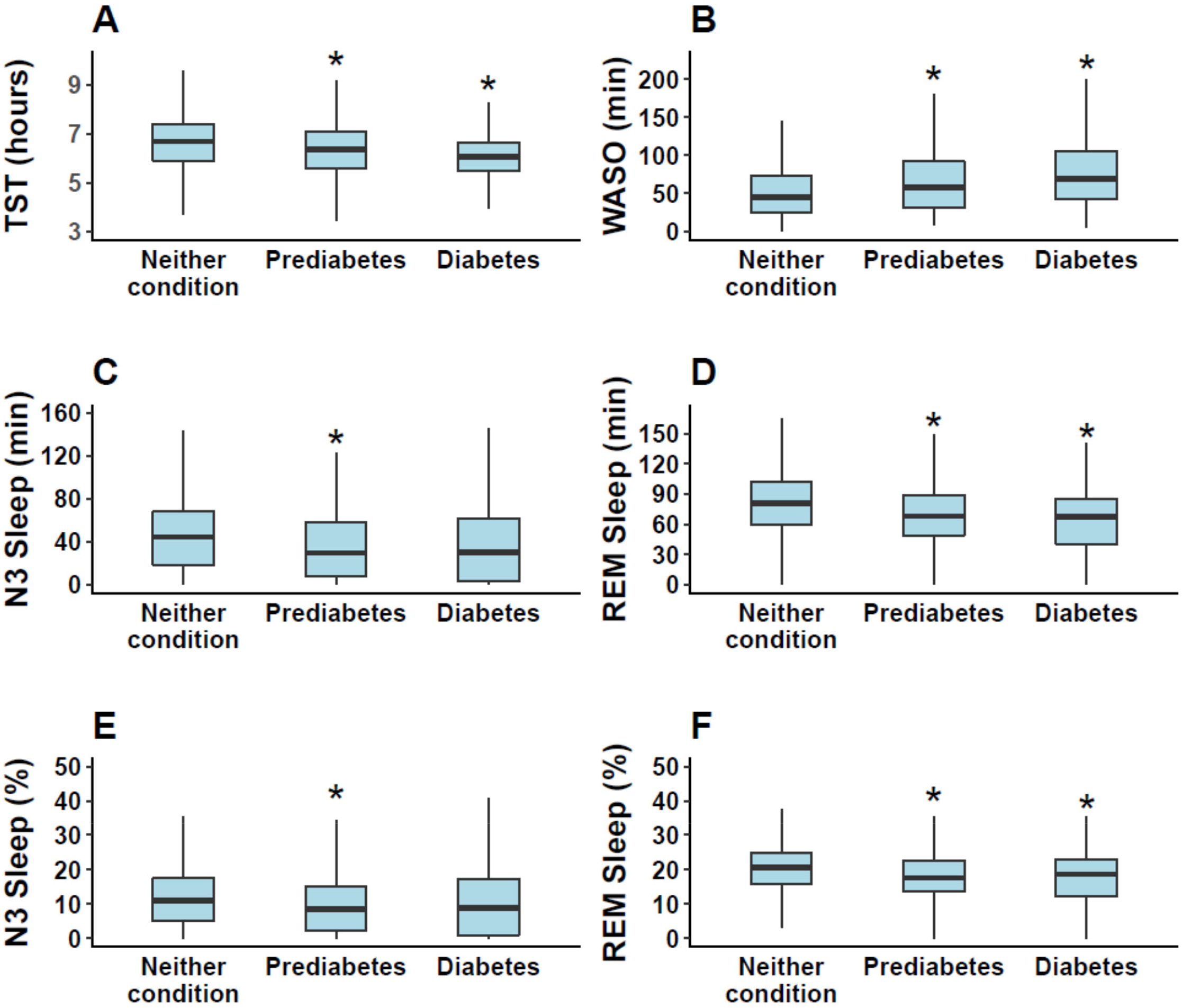
Comparison of the distribution of PSG measures among individuals with neither condition, prediabetes and diabetes with AHI<30. Asterisk denotes significant unadjusted differences (p<0.05) between neither condition and either prediabetes or diabetes group.

In regression models adjusted for age, gender, BMI and AHI (**Table 2**), participants with diabetes had shorter TST (−14 minutes on average). Also, both participants with diabetes and participants with prediabetes averaged approximately 6-7 minutes less REM compared to those with neither condition. People with prediabetes also had lower REM percentage (−1.4%) than those with neither condition, although people with diabetes did not differ. Finally, in the fully adjusted models, people with diabetes had longer average N3 duration (+7.6 min) and higher N3 percentage (+2.4%) than those with neither condition. There were no differences in WASO among the three groups.

We also conducted secondary analyses to exclude participants with moderate sleep apnea (AHI ≥ 15). The unadjusted comparisons of the PSG characteristics are depicted in **Figure 2** and the regression results are presented in Table 3. The results were similar to the primary analyses. Diabetes was associated with lower TST and higher N3 duration and percentage, and prediabetes was associated with lower REM duration and percentage. Finally, we conducted sensitivity analyses to investigate effect modification by age, but there was no significant effect modification.

**Figure 2:**
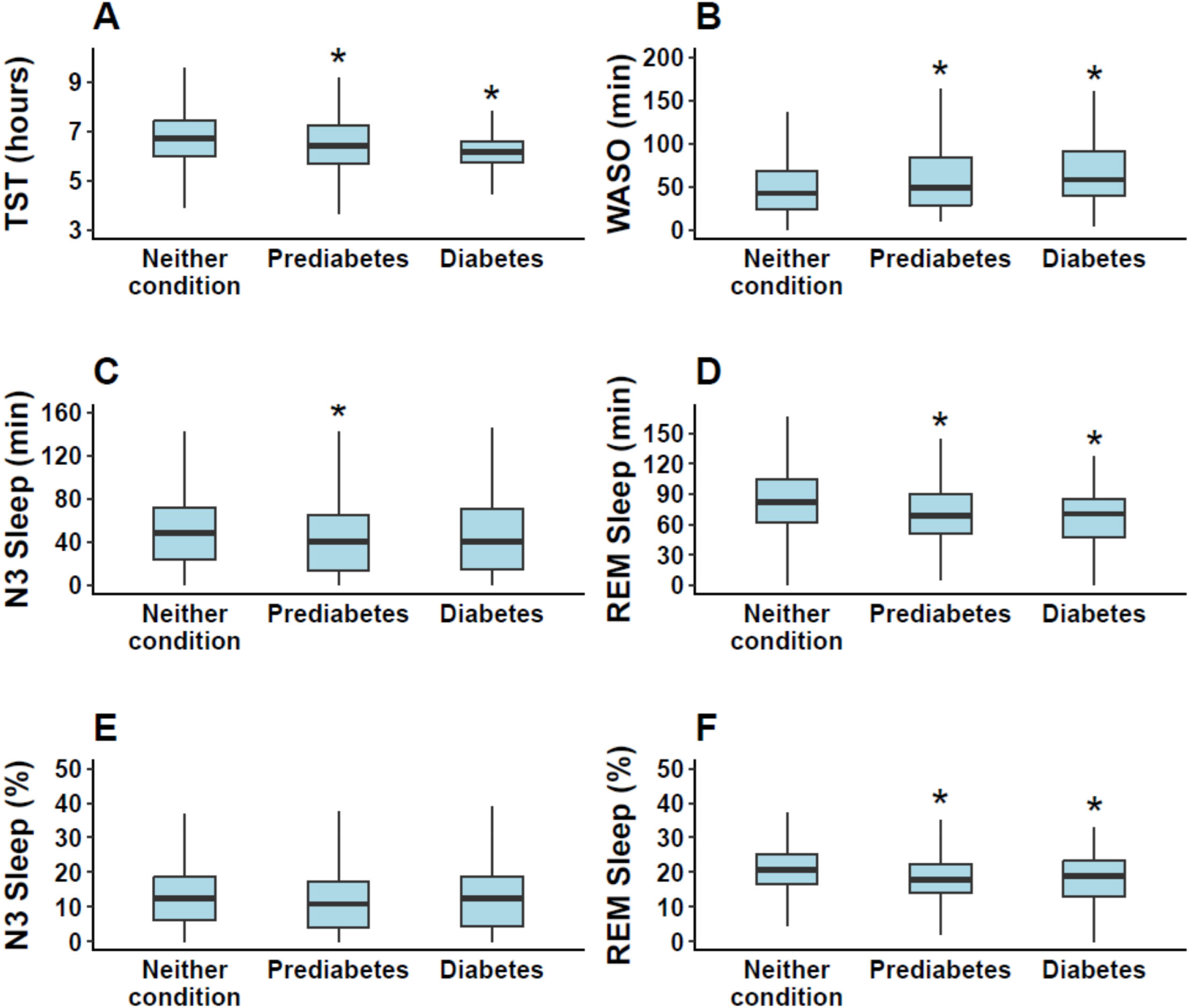
Comparison of the distribution of PSG measures among individuals with neither condition, prediabetes and diabetes with AHI<15. Asterisk denotes significant unadjusted differences (p<0.05) between neither condition and either prediabetes or diabetes group.

**Table 3.** Regression models comparing PSG measures among individuals with neither condition, prediabetes and diabetes adjusting for covariates in sample with AHI<15 events/hour.

|  |  | Unadjusted Model | Adjusted Model<sup>a</sup> |
| --- | --- | --- | --- |
| PSG Measure | Diabetes Group | Beta (95% CI) | Beta (95% CI) |
| TST (h) | Neither condition | referent | referent |
|  | Prediabetes | -0.22 (-0.39, -0.06) | -0.05 (-0.22, 0.13) |
|  | Diabetes | -0.49 (-0.73, -0.25) | -0.28 (-0.53, -0.02) |
| WASO (min) | Neither condition | referent | referent |
|  | Prediabetes | 10.1 (3.7, 16.4) | -1.6 (-8.2, 5.0) |
|  | Diabetes | 16.1 (6.7, 25.5) | 2.7 (-6.9, 12.2) |
| N3 duration (min) | Neither condition | referent | referent |
|  | Prediabetes | -6.7 (-12.3, -1.1) | 2.7 (-3.1, 8.4) |
|  | Diabetes | -2.7 (-11.0, 5.6) | 8.7 (0.3, 17.0) |
| REM duration (min) | Neither condition | referent | referent |
|  | Prediabetes | -13.8 (-18.6, -8.9) | -6.2 (-11.3, -1.0) |
|  | Diabetes | -16.4 (-23.6, -9.2) | -7.0 (-14.5, 0.5) |
| N3 % | Neither<br>condition | referent | referent |
|  | Prediabetes | -1.3 (-2.7, 0.1) | 0.8 (-0.7, 2.3) |
|  | Diabetes | 0.3 (-1.8, 2.4) | 2.8 (0.6, 4.9) |
| REM % | Neither<br>condition | referent | referent |
|  | Prediabetes | -2.7 (-3.7, -1.6) | -1.3 (-2.4, -0.2) |
|  | Diabetes | -2.5 (-4.0, -0.9) | -0.8 (-2.4, 0.8) |

<sup>a</sup> Model adjusted for: age, gender, BMI, AHI

## Conclusions

Our goal was to compare sleep architecture in people with diabetes or prediabetes to those with neither condition after removing potential confounding due to sleep-disordered breathing. Our hypothesis was only partially supported by the analyses. Consistent with our hypothesis, both participants with prediabetes and diabetes had less REM sleep compared to those with neither condition. This association persisted even after accounting for potential confounders of this association, including age, gender and BMI. However, contrary to our hypotheses, we observed a longer duration of N3 sleep in participants with diabetes after adjusting for covariates. The scientific premise of this report was based on two general findings. First, experimental sleep restriction or N3 suppression can impair glucose metabolism or alter energy intake. Second, prior work has demonstrated greater sleep disturbances, particularly sleep-disordered breathing, among people with diabetes. Our finding that prevalent diabetes and prediabetes were associated with less REM sleep is consistent with prior work. With over 1,000 participants in our sample, ours is one of the largest studies to investigate this relationship. One other large study (13), also reported a decrease in REM percentage among individuals with type 2 diabetes (they did not examine prediabetes). Other smaller studies vary in their findings when comparing those with diabetes to those without; two found no differences in REM sleep (14; 16) while another (n=22 in each group) found a higher REM percentage in people with type 2 diabetes (15). Our finding that N3 duration was greater among people with diabetes was contrary to other studies that either found less N3 among those with diabetes (16) or no difference (13; 14). The discrepancies in findings among these studies, including ours, may be due to smaller sample sizes in most other studies, demographic differences between studies, including age, differences in covariate adjustment and our exclusion of those with moderate and severe sleep disordered breathing.

There are many potential physiological mechanisms linking diabetes to less REM sleep, beyond the presence of sleep disordered breathing. One potential mechanism is inflammation. It is well-known that diabetes and obesity, which is common in diabetes and prediabetes, cause chronic inflammation (21), and inflammatory markers have been associated with greater REM latency (time between sleep onset and first REM epoch) (22), although the latter study did not observe a significant association between inflammation and REM percentage. Another study examined several inflammatory markers and PSG characteristics and found that greater REM percentage was associated with higher levels of complements but also lower levels of IL-6 and CFI (23), all of which are proinflammatory. Therefore, the association between sleep stages and inflammation, particularly in the setting of diabetes, requires further investigation. Diabetes also causes a multitude of vascular complications, and microvascular complications in diabetes have been associated with decreased REM duration and lower sleep efficiency (24). However, since participants with prediabetes also have less REM sleep, it is less likely that the difference in sleep architecture is due to cerebrovascular or atherosclerotic differences. Another common clinical sequela of diabetes is peripheral neuropathy, and neuropathic pain has a strong association with sleep disturbance (25). We did not have measures of neuropathy or pain in this study, so we could not explore these associations.

The finding that diabetes is associated with greater N3 duration was unexpected. One possible explanation for this finding is also related to inflammation, as some of the studies discussed above also found that greater levels of proinflammatory makers are associated with higher N3 (22; 23). Additionally, if people with diabetes are chronically sleep deprived (as suggested by their lower total sleep time), they may have more N3 and less REM due to the homeostatic regulation of sleep. It is well known that N3 sleep is homeostatically controlled, with increased sleep deprivation and associated increased sleep pressure leading to more N3 sleep (26). Further, it may be possible that sleep need is altered in diabetes resulting in greater sleep pressure as well. Finally, the temporal distribution of sleep stages across the night may be somewhat altered in diabetes. Normally, N3 occurs primarily in the first half of the night, while REM occurs primarily in the second half of the night. Nonetheless, there are periods of overlap and it is possible that N3 is prioritized over REM at these times, resulting in both higher N3 and lower REM. Understanding whether these alterations in sleep architecture have implications for disease management is an important next step.

This study had several strengths, which included a large sample size, full PSG recordings, as well as our examination of individuals with prediabetes. Our study is the first to our knowledge to observe poorer sleep architecture (i.e., less REM) among people with prediabetes. There are some important limitations of our study as well. These include the cross-sectional study design, which does not allow us to determine whether sleep architecture changes after the development of prediabetes or diabetes. In addition, we do not know the duration of disease for each person. Finally, the use of diabetic medication was self-reported, which raises the possibility that medications were under- or over-reported.

Since sleep architecture has been linked to impaired glucose homeostasis, it is important to understand how these sleep stages differ among people with type 2 diabetes or prediabetes. We found that diabetes and prediabetes are both associated with less REM sleep, even in the absence of moderate-severe sleep apnea. Future work should determine whether these differences, i.e. less REM sleep or more N3, is associated with glucose control, or other health outcomes, in these patients. Similarly, the association with N3 should be examined further to see if it is associated with disease management or habitual sleep patterns. Sleep health, including sleep architecture, may play an important role in the health and well-being of patients with type 2 diabetes or prediabetes and warrants further consideration.

## Data Availability

All data produced in the present study are available upon reasonable request to the authors

## Funding and Assistance

This work was supported by NIH grant 1R01HL141881.

## Author Contributions

D.M.C. designed the study, drafted and reviewed the manuscript. T.P.T. designed the study, drafted and reviewed the manuscript. S.J.A. and D.A.A. drafted and reviewed the manuscript and analyzed data. F.B., M.v.S., and A.P. designed the study, acquired data, and reviewed the manuscript. K.L.K. conceived of and designed the study, analyzed data, drafted and reviewed the manuscript, obtained funding, provided administrative support, and supervised the study. K.L.K. is the guarantor of this work, and as such, had full access to all the data in the study and takes responsibility for the integrity of the data and the accuracy of the data analysis.

## Prior Presentation

A non-peer-reviewed version of this article was presented at the Advances in Sleep and Circadian Science conference on February 18, 2023.

